# Earlier timing of seasonal respiratory infections and seasonal mortality during the COVID-19 pandemic

**DOI:** 10.1101/2024.12.17.24319104

**Authors:** Michael Sieber, Arne Traulsen

## Abstract

Seasonal respiratory infections typically surge within a limited time window, but the exact timing within a given year is hard to predict. The disruptions caused by the COVID-19 pandemic led to dramatic changes in the transmission dynamics of many pathogens, providing a unique opportunity to study the determinants and robustness of the seasonal timing of epidemics. Combining detailed data on acute respiratory infections from Germany with an epidemiological model, we analyzed changes in the timing of seasonal epidemics. The seasonal surge in infections occurred substantially earlier during the COVID-19 pandemic, and was reflected in a corresponding shift in the seasonality of all-cause mortality. We show that this is a consistent, but transient outcome of disrupted epidemic seasonality, predictable from basic epidemiological principles.

## Introduction

Many infectious respiratory diseases in human populations experience a seasonal variation in transmission, leading to recurring epidemics (*1–5*). The non-pharmaceutical interventions (NPIs) implemented during the COVID-19 pandemic constituted a major disruption of the seasonality for many endemic respiratory diseases, resulting in wide-spread changes to their usual seasonal dynamics (*6–11*). One of the most striking examples of this effect was the almost complete disappearance of influenza and respiratory syncytial virus (RSV) for one season or even longer (*12–15*).

It had been suggested during the COVID-19 pandemic that such disruptions can lead to changes in the severity and timing of the seasonal dynamics of the respective diseases (*16–20*). Out-of-season waves of respiratory infections following COVID-19-associated NPIs have subsequentely been reported from around the world (*13, 15, 21–30*), which were not necessarily more severe than previous seasons (*31*).

Whether these unusual epidemic patterns are a predictable consequence of disruptions of seasonal infection dynamics and also translate to changes in temporal patterns of mortality has not been systematically evaluated. While the major drivers of disease seasonality such as changes in the abiotic environment, vector seasonality and seasonal changes in host behaviour (*1, 32, 33*) determine the overall seasonality of diseases, the determinants of the exact timing remain largely unknown (*1*).

With more than one full seasonal cycle having passed since the Public Health Emergency of International Concern due to COVID-19 has been declared over in May 2023, a more complete picture of the magnitude and duration of the disruptive changes of seasonal epidemics due to NPIs emerges. The interplay of an emerging respiratory disease and the disruption and resurgence of already circulating diseases provides a unique opportunity to study the drivers of seasonal forcing of respiratory diseases and the factors determining the seasonal timing of epidemics (*1*).

## Transient shift in timing of respiratory disease season

We analyzed all available data for weekly incidences of self-reported symptomatic respiratory infections, doctor’s visits for acute respiratory infections (ARI) and hospitalisations due to severe acute respiratory infections (SARI) in Germany for the last 10-12 years. The incidences of respiratory infections follow a clear seasonal pattern, increasing four to five-fold in late fall and winter compared to the summer months (Figs. 1a-c). This seasonal pattern was remarkably stable before the COVID-19 pandemic, with incidences consistently peaking during just a few weeks in February and March each year (Figs. 2a-c).

**Figure 1.**
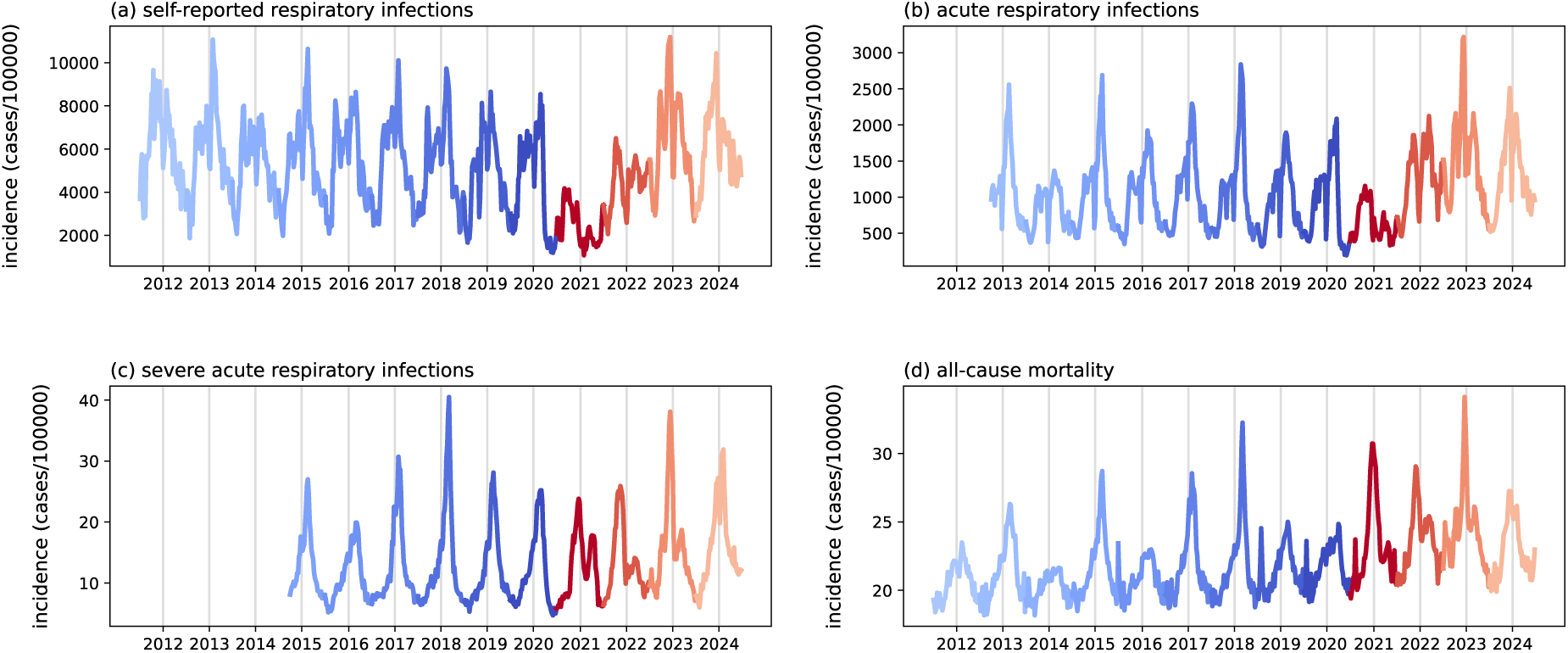
Timeseries data showing the strong seasonal component of respiratory infections and all-cause mortality in Germany. Pre-pandemic seasons are shown in blue and (post-)pandemic seasons in red colors. (a) Weekly incidence of self-reported, symptomatic respiratory infections (SRI). (b) Weekly incidence of acute respiratory infections (ARI). (c) Weekly incidence of hospitalized severe acute respiratory infections (SARI). (d) All-cause mortality as number of deaths per 100.000 per week. The winter peaks are often interspersed with a sharp summer peak, corresponding to heat-related mortality in particularly hot summers.

**Figure 2.**
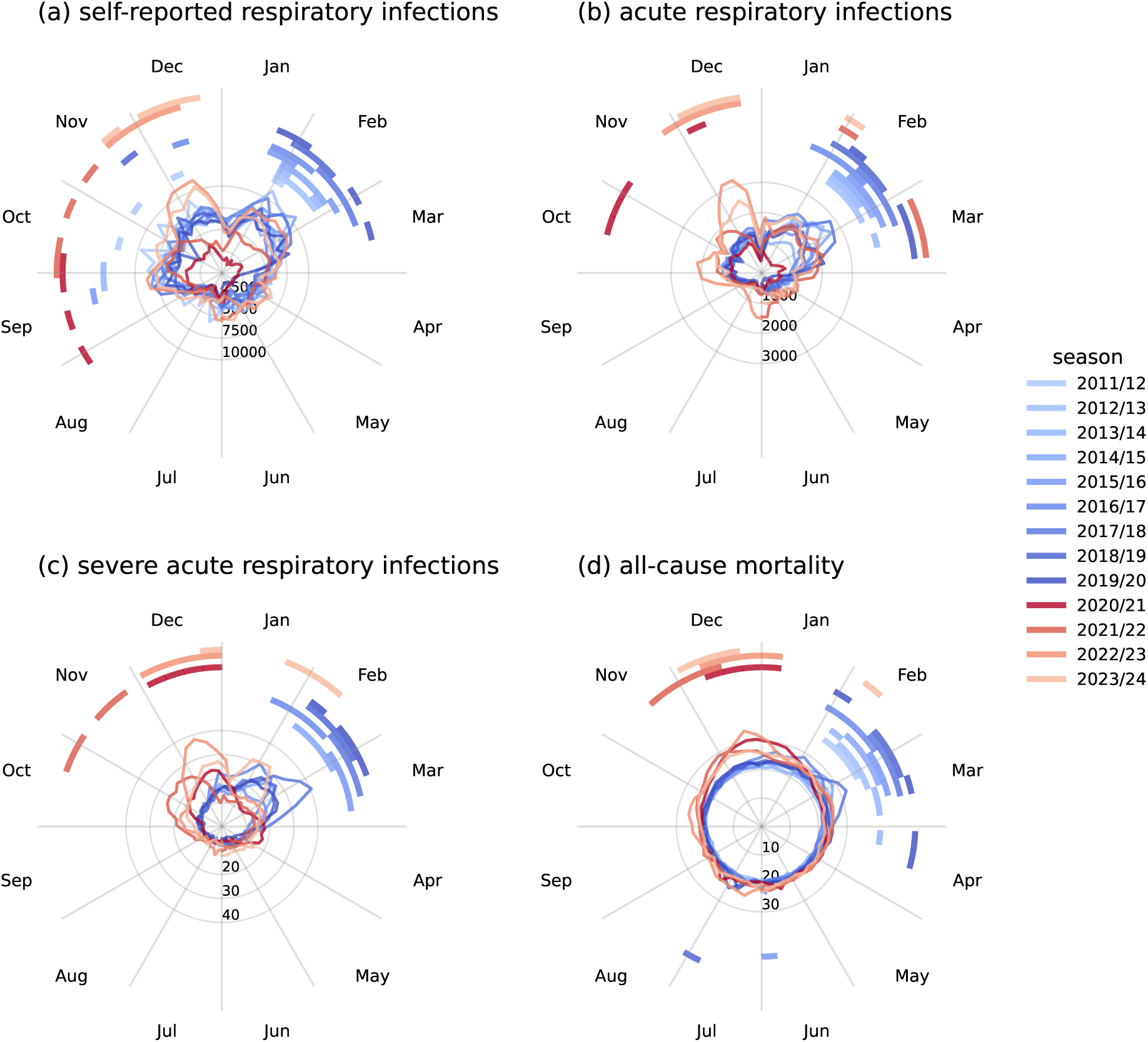
The same data as in Fig. 1 in a polar plot to highlight the shift in the timing of seasonal epidemics. As in Fig. 1, pre-pandemic seasons are shown in blue and (post-)pandemic seasons in red. The bars above the timeseries indicate the four weeks with the highest incidences in each season. Before the pandemic, those peak weeks occurred almost exclusively in February and March. During and after the pandemic they were shifted to December or even earlier. (a) Weekly incidence of self-reported, symptomatic respiratory infections (SRI). (b) Weekly incidence of acute respiratory infections (ARI). (c) Weekly incidence of hospitalized severe acute respiratory infections (SARI). (d) All-cause mortality as number of deaths per 100.000 per week.

The NPIs implemented in 2020/21 during the first winter of the the COVID-19 pandemic in Germany resulted in a reduced transmission not only of Sars-CoV-2, but also of many other respiratory infections. For example, seasonal influenza and RSV were almost completely absent in the winter of 2020/21 (Fig. 3). This reduced transmission of respiratory pathogens due to NPIs is reflected in a reduced number of self-reported respiratory infections and doctor’s visits for ARIs in the fall and winter of 2020/21 (Fig. 1a,b). This reduction was found across all age groups, with weekly incidences reduced by 50% or more compared to previous seasons (Figs. S1b, S2b, S3b). The effect was even more striking for hospitalisations of 0-4 years olds, with no discernible seasonal increase of SARI cases at all in the winter of 2020/21 (Figs S1c).

**Figure 3.**
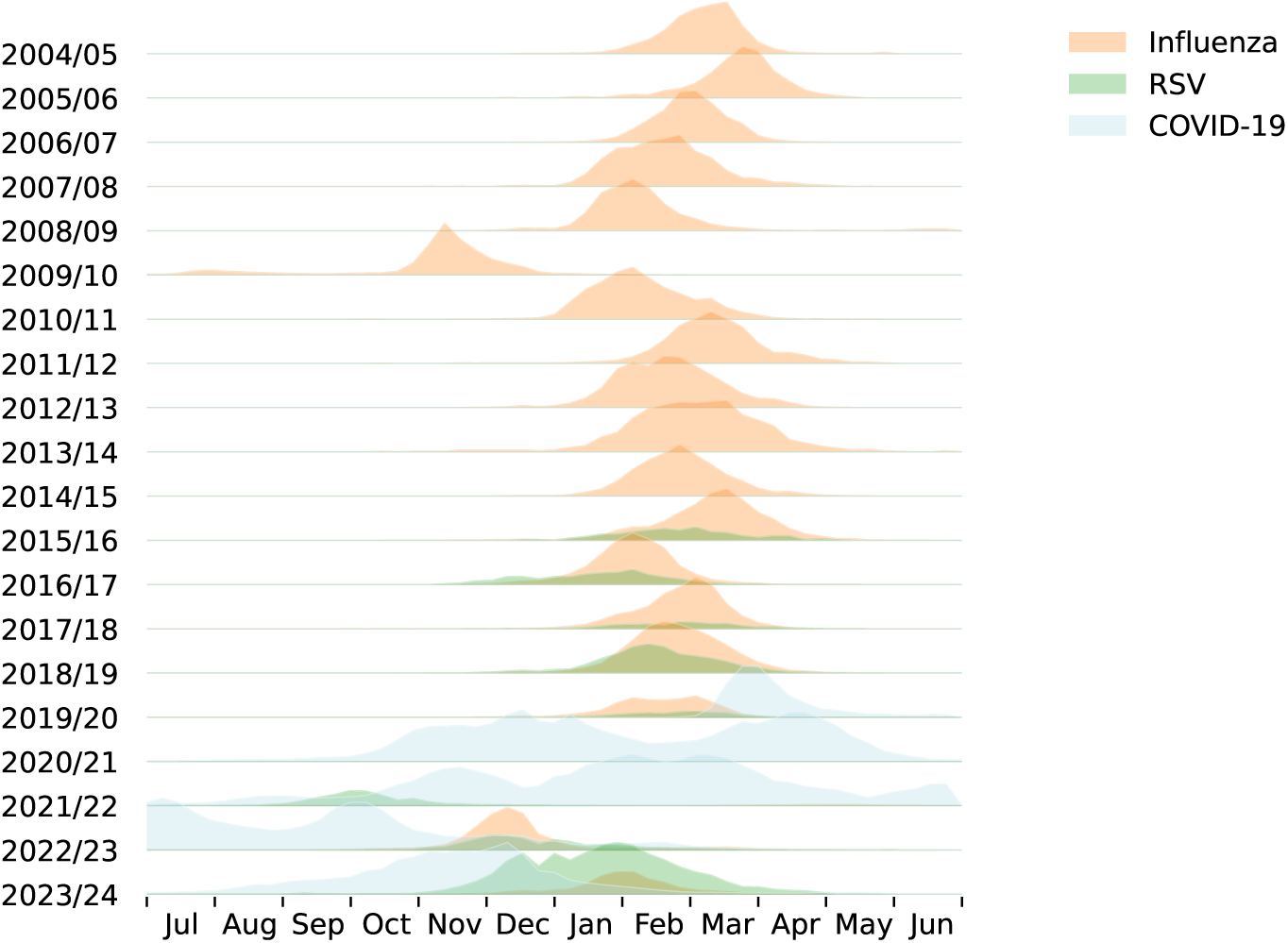
Normalized weekly incidences of laboratory confirmed COVID, seasonal influenza and RSV in Germany. Influenza and RSV very consistently peaked in February and March, except during the swine flu pandemic 2009/10 and since the onset of the COVID pandemic. For RSV we observe a gradual shift of the peak back to its normal timing during the last three seasons, although the current RSV season still started much earlier than usual. After the early peak of the first post-pandemic influenza season 2022/23, the current 2023/24 influenza season has already moved almost back to its pre-pandemic timing.

Despite this anomalous season at the start of the COVID-19 pandemic, the overall epidemiological dynamics of ARI cases continued to be dominated by a clear seasonal signal (Fig. 1a-c), in particular for the severe cases that required hospitalisation (Fig. 1c). At the same time, rapid evolution of SARS-CoV-2 led to out-of-season waves of emerging viral variants, most prominently the spread of the Alpha variant from January to April 2021, and the Omicron variant from January to March 2022. These waves appear as overlaying the dominating seasonal pattern (Fig. 1a-c).

While the overall seasonality of acute respiratory diseases appeared unchanged after the anomalous 2020/21 season, there was a striking shift in timing: In the first winter of the COVID-19 pandemic in 2020/21 and the two following seasons from 2021/22 to 2022/23, the onset and peak of respiratory disease season was shifted by several weeks, each occurring 8-12 weeks earlier than in all of the observed previous seasons (Figs. 2a-c).

For the 2020/21 winter, this shift was driven by the seasonally early epidemic spread of COVID-19 (Fig. 3), which was responsible for the majority of ARI and SARI cases in the absence of influenza and RSV. After being absent in 2020/21, RSV re-emerged in September of 2021/22, very early in the season and almost simultaneously with the start of the next winter wave of COVID-19 (Fig. 3). The COVID-19 pandemic was at this time mainly driven by the highly pathogenic Delta variant (Fig. S4).

As a consequence the incidence of hospitalized SARI cases already peaked in November, earlier than in any other observed season and 3-4 months before the usual pre-pandemic timing (Fig. 2c). For the ARI cases the 2021/22 season is more complicated, as the highly transmissible, but less pathogenic Omicron variant began to rapidly replace the Delta variant in late 2021 and early 2022 (Fig. S4). This emerging variant lead to a peak in ARI cases in March 2022, which did not lead to a corresponding peak in hospitalisations, presumably due to the generally lower pathogenicity of the Omicron variant. In 2022/23 seasonal influenza, re-emerging after being almost completely absent in 2020/21 and 2021/22, had an early epidemic peak in December, 2-3 months before its usual peak in February or March (Fig. 3). Simultaneously with influenza in 2022/23 the RSV epidemic occurred, two months later than the very early season in 2021/22 right after its re-emergence, but still two months earlier than usual (Fig. 3).

This suggests a shift back towards the usual, pre-pandemic seasonal timing for RSV, a pattern that was confirmed for both RSV and influenza in the most recent season in 2023/24. A similar shift back to pre-pandemic seasonal patterns had also been reported in the US (*28*). The 2023/24 season was characterized by a succession of COVID-19 early in the season from September to December, followed by seasonal influenza and RSV, which both peaked almost back at their normal timing in late January and early February (Fig. 3). This suggests that the shift in seasonal timing for previously circulating diseases in Germany was transient and was gradually reversed within one or two seasons.

## Corresponding shift in seasonality of mortality

The excess mortality associated with the COVID-19 pandemic has received a lot of attention, showing that the number of deaths was substantially higher than expected around the world (*34*). But changes to the temporal dynamics of mortality have received relatively little systematic attention. For this, we analyzed the timeseries of weekly all-cause mortality in Germany (*35*), which also follows a clear seasonal pattern (Fig. 1d and 2d).

In Germany, weekly mortality is correlated with the weekly incidence of ARI and SARI cases, generally increasing in fall and peaking in late winter (Fig. 2d). While peak mortality can vary by more than 30% season-by-season, the timing of the seasonal peak in mortality was remarkably stable before the COVID-19 pandemic. Mortality generally peaked in February or March, coinciding with the pre-pandemic seasonal peaks in ARI and SARI cases (Fig. 2d). Naturally, the dynamics of all-cause mortality are dominated by the older age groups, and this clear seasonal pattern is absent from the youngest age groups (Fig. S1d).

Strikingly, during and after the COVID-19 pandemic the seasonal peak in mortality occurred 2-3 months earlier, in December and early January (Fig. 2d). This is a shift very similar to the one observed for respiratory infections and a strong signal of the effect the pandemic had not only on epidemiological processes, but also seasonal dynamics of mortality. The close match between the shift in mortality and the shift in the timing of respiratory infections suggests that most of the seasonal component of mortality can in fact be attributed to respiratory infections and their direct and indirect side effects.

## Shift in seasonal timing in an SIRS model

The transient shift in seasonal timing of respiratory diseases can be understood within an established SIRS (susceptible-infectious-recovered-susceptible) model with seasonal forcing. Such SIRS or related SEIR (susceptible-exposed-infectious-recovered) models have been used to shed light on the epidemiological dynamics of seasonal diseases, including sudden transitions between different epidemiological patterns (*36*), and the transient and long-term effects of perturbations of the seasonal forcing (*37*).

In the simplest case, the SIRS model describes the spread of an infectious disease in a population of constant size with the fraction of susceptible individuals *S*, the fraction of infected individuals *I*, and the fraction of recovered and immune individuals *R*, so that *S* + *I* + *R* = 1. Susceptible individuals get infected with the intrinsic transmission rate *β* (*t*) by infected individuals. The transmission rate varies with time, reflecting that seasonally changing environmental conditions influence the transmissibility of the disease. We assume a periodically driven transmission rate

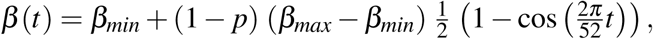

which for *p* = 0 oscillates between a minimal transmission rate *β_min_* and a maximum transmission rate *β_max_*. The factor 1 − *p* in this model reflects the reduction of the amplitude of the seasonal forcing due to non-pharmaceutical interventions (NPIs), so that *p* = 0 during a usual season in the absence of NPIs and 0 *< p* ≤ 1 during periods of NPIs. We assume that NPIs can not reduce transmission below the minimal rate *β_min_*even at 100% efficacy, but our results do not depend on the specific way NPI efficacy *p* affects transmission. The period of the seasonal forcing is chosen as 52 weeks, with the maximum transmission rate occurring at *t* = 26 weeks, defined as the turn of the calendar year and mid-season.

We fix the minimal transmission rate at *β_min_* = 0.6 and the maximum transmission rate at *β_max_* = 3. After infection an individual remains infective for a duration of *ρ*^−1^ = 1 week, after which it recovers and enters the fraction *R*. Recovered individuals are protected against reinfection for a duration of immunity of *ω*^−1^ = 40 weeks. Waning immunity is the result of a complex interaction between the host’s immune system and antigenic variation of the causal pathogen, and the exact duration of protection is uncertain for many respiratory diseases (*38*). This model leads to annual epidemics and has previously been used to describe the spread of seasonal influenza (*37*), and with similar parameters for RSV (*39, 40*). Unless stated otherwise, NPIs reduce transmission by 30% (*p* = 0.3).

The dynamical equations for this SIRS model are

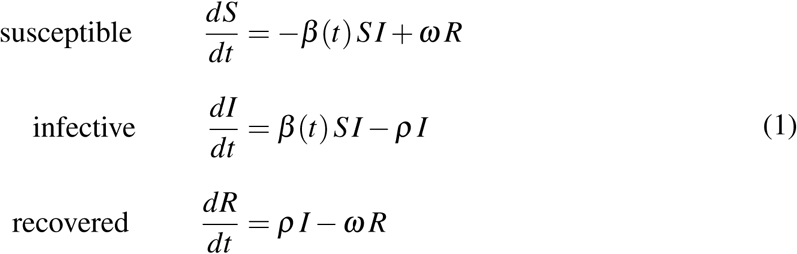

A timeseries obtained from the numerical solution of the model is presented in Fig. 4a, starting from an initially completely susceptible population. This corresponds to the scenario of a novel disease spreading in a population without pre-existing immunity, as for example at the onset of the COVID-19 pandemic. After introduction of the disease the seasonally varying transmission rate leads to seasonal epidemics with a very consistent timing (Fig. 4b). For the chosen parameter values the peaks of the recurring epidemics are reached in a narrow time window just after the transmission rate has reached its seasonal maximum.

**Figure 4.**
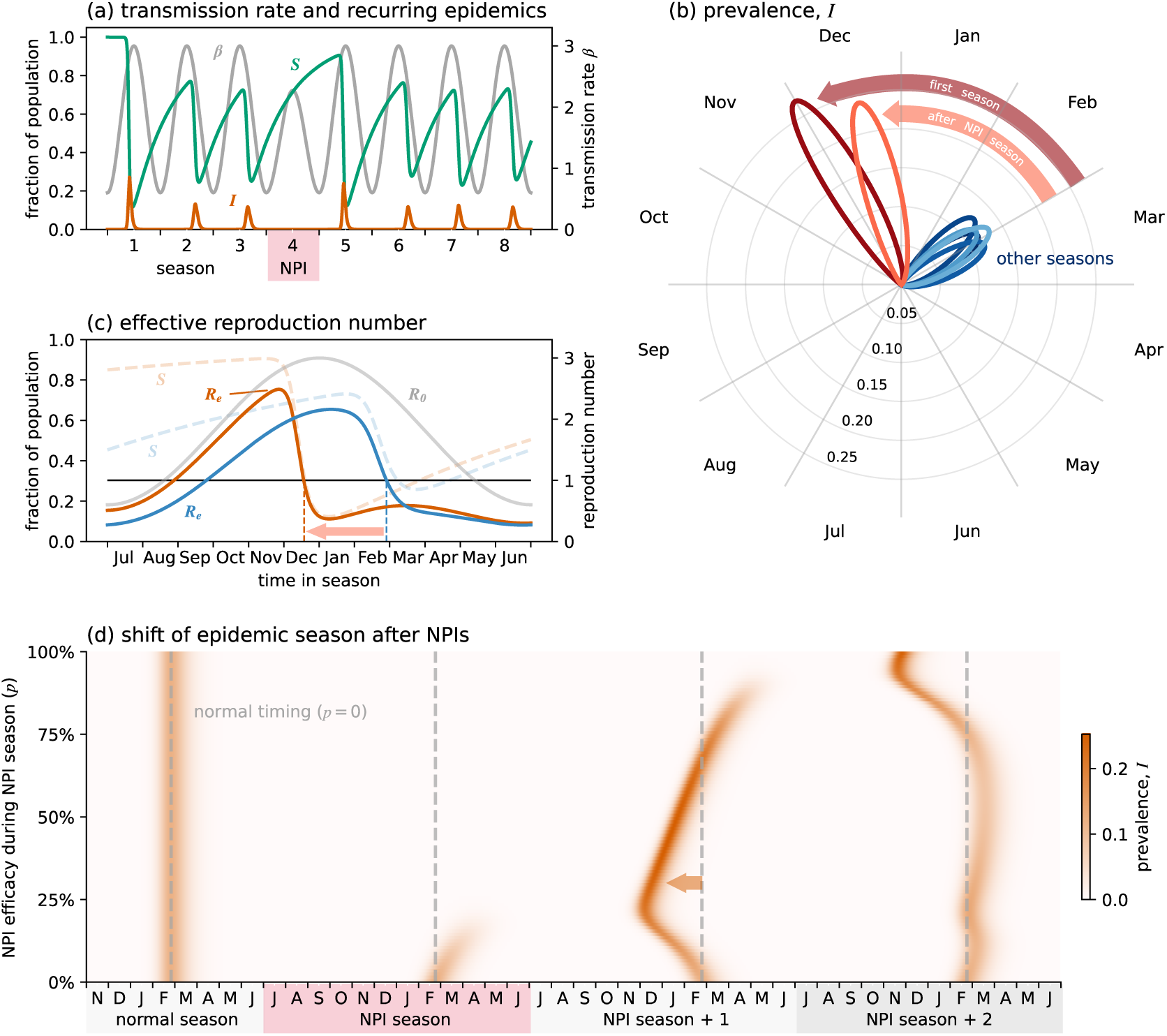
(a) Timeseries obtained from model (1). During the NPI season marked in red the maximum transmission rate *β_max_* is reduced by 30%. This results in the suppression of the seasonal epidemic and an earlier epidemic in the following season. (b) Same data as in (a) in a polar plot, highlighting the shift in peak timing. The seasonal epidemics after the initial introduction of the disease and right after the NPI season happen earlier than during other seasons. (c) The effective reproduction number *R_e_* is higher earlier in the season after a prolonged buildup of susceptibles *S* due to NPIs (orange lines), which is similar to the scenario of a newly arising disease with limited pre-existing population-level immunity. The blue lines show the situation during a usual season without disruptions. The dashed vertical lines mark the peak of the seasonal epidemic in the respective seasons, showing the shift forward after an NPI season. (d) Effect of different levels of NPI stringency on the shift of subsequent seasonal epidemics. The NPIs generally lead to earlier epidemics in the following season, unless NPIs are so effective that they almost eradicate the disease. In this case the following season can be slightly later and weaker than usual, or even be entirely skipped (here for *p >* ca. 75%). The disease resurgence two seasons later is then again earlier than usual.

The very first epidemic is an exception to this pattern, as it starts much earlier in the seasonal cycle and reaches its maximum before maximum transmissibility has been reached at the turn of the year (Fig. 4b). This can be understood in terms of the number of secondary infections caused by one infective individual, which in a completely susceptible population (*S* ≈ 1) is determined by the basic reproduction number *R*_0_ = *β* (*t*)*/ρ*. This number is directly proportional to the intrinsic transmissibility *β* (*t*) of the disease, and thus it changes with the time of season. But this is only valid in a fully susceptible population and more generally the reproduction number not only depends on the transmission rate *β* (*t*), but also on the current size of the fraction *S* of the population that is susceptible. This leads to the effective reproduction number *R_e_* = *R*_0_ *S*, where both the basic reproduction number *R*_0_ and the susceptible fraction *S* change dynamically in time.

The disease will only spread and cause an epidemic if *R_e_>* 1, which in the model (1) exactly corresponds to a positive growth rate of the infective fraction *I*. In this case, the size of the susceptible fraction *S* decreases as more individuals become infected and eventually recover, until *R_e_* falls below 1 again. At this point the epidemic has reached its seasonal peak and the infective fraction starts to decrease. After some time, the susceptible fraction will start to increase again due to waning immunity until the cycle begins again.

Since outside of NPI-periods the transmission rate *β* has fixed temporal dynamics, it is the dynamic size of the susceptible fraction *S* that determines the exact timing of when the effective reproduction number will be above or below 1. Generally, the larger the fraction *S* is earlier in the seasonal cycle, the earlier the effective reproduction number will cross this threshold and the earlier the epidemic will start relative to its underlying seasonal forcing. After introduction of a disease into a fully susceptible population (*S* ≈ 1) the effective reproduction number *R_e_* is close to the basic reproduction number *R*_0_ = *β* (*t*)*/ρ*, and consequentely the epidemic starts at the earliest possible time, i.e. as soon as *β* (*t*)*/ρ >* 1 (Fig. 4c).

In the subsequent seasons, when there is some amount of pre-existing immunity on the population level (*S <* 1), the effective reproduction number *R_e_ < R*_0_ governs the infection dynamics and thus the epidemic will start and peak later in the seasonal cycle (Fig. 4c). Due to the constant rate of immune waning *ω* and in the absence of other disruptions, the increase of susceptibles between two consecutive epidemic peaks follows a very consistent pattern. This results in subsequent epidemics having a very consistent timing later in the seasonal cycle compared to the initial season (Fig. 4b).

If an epidemic season is reduced or entirely skipped due to NPIs, this consistent temporal pattern is disrupted. After an NPI period and a skipped seasonal epidemic, the buildup of susceptibles in the population due to prolonged immune waning leads to a higher *R_e_* earlier in the season (Fig. 4c). This in turn results in an earlier onset of the corresponding seasonal epidemic, which will generally fall somewhere between the timing of the very first epidemic season and the usual, later timing of unperturbed seasons (Fig. 4b).

The extent of the shift away from the usual seasonal timing depends on the size of the susceptible fraction reached during and after the NPI period. This buildup of susceptibles ultimately depends on the ability of the NPIs to suppress the disease, in the model described by the efficacy *p* of NPIs. Varying the efficacy *p* of the NPIs shows that the seasonal epidemic is effectively contained if the transmission rate is reduced by 20% or more (Fig. 4d). At the same time the following season is shifted forward towards an earlier timing. For the chosen parametrization this shift is most pronounced for an NPI efficacy of ca. 25%, but the forward-shift is observed for the larger part of the efficacy range up to *p* ≈ 70% (Fig. 4d). Above *p* ≈ 70%, the seasonal epidemic is slightly delayed, despite the effective reproduction number *R_e_* becoming larger than one earlier in the season.

This delay arises because with very effective NPIs the prevalance *I* drops so low that the exponential growth rate of the disease is initially so low that a noticable epidemic develops only later in the season. If the NPIs are so effective that the disease is almost driven to extinction (*p >* 90%), the exponential growth rate is in fact so low that there is not enough time for the infection to grow into an epidemic within the seasonal window. In this case the epidemic in the first season after the NPIs is skipped entirely and the disease re-emerges in the second season after the NPIs. This resurgence then again happens much earlier compared to the usual epidemic timing (Fig. 4d). This delay or skipping of an additional season after very effective NPIs can be reduced or even reversed if there is a small, constant influx of infecteds, which prevents the prevalence *I* from dropping to extremely low values.

The exact size and timing of seasonal disease outbreaks also depends on the dynamics of the underlying seasonal forcing. But the general pattern of diseases spreading earlier in their seasonal cycles either directly after introduction or after reduced transmission in a previous season is generally robust to variations in the minimum and maximum transmission rates (Fig. S5). If, however, maximum transmission rates are very high the usual seasonal peaks already happen relatively early, and disruptions by NPIs have no potential to move them much further forward (Fig. S5e).

Another result from this simple model is that if waning of immunity happens on a much faster time scale compared to the seasonal cycle (*ω*^−1^ ≪ 52 weeks), a similar shift of the epidemic timing following a perturbation of the seasonal forcing would not be observed (Fig. S7). In this case the population would quickly be made up mostly of susceptibles again following an epidemic, and the following season would thus again start at the earliest possible time, solely determined by the intrinsic transmission rate *β* (*t*).

## Discussion

In general, two factors determine if and when seasonal epidemics occurr: (i) seasonal variation of pathogen transmission driven mainly by environmental factors and (ii) the fraction of the population that is susceptible (*1*). But the relative importance of these factors and how they work together to determine the exact timing is not clear. The NPIs associated with the COVID-19 pandemic and the resulting disruption of the pre-pandemic seasonal pattern has allowed us to disentangle these factors in more detail.

We observed a significant shift in the timing of seasonal respiratory disease epidemics in Germany during the fall and winter of 2021/22 and 2022/23. A corresponding shift is also observed in the seasonal pattern of all-cause mortality. Together with the complete absence of the most important respiratory diseases during the 2020/21 and 2012/22 seasons, this shift is one of the clearest signals of the impact that the developing SARS-CoV-2 pandemic and associated mitigation measures had on established seasonal epidemics.

This shift is even more profound considering that the timing of seasonal epidemics in Germany has been remarkably consistent, with highest incidences typically occurring within a relatively narrow time window in February and March. In fact, in the last 20 years there is only one precedent of a similar seasonal shift, caused by the novel 2009 H1N1 influenza which lead to a global pandemic (*41*). This early-onset influenza epidemic also led to a transient change in the timing of seasonal epidemics of RSV and other respiratory diseases. In this case the change in timing was not as pronounced and not necessarily all in the same direction (*41*), which may be more a result of direct viral interference rather than the more limited implementation of NPIs during this pandemic (*42*).

Supplementing our analysis of respiratory disease epidemics in Germany with an SIRS model has enabled us to show that seasonal variation of transmission creates the necessary window of opportunity for epidemics, but it is the availability of susceptibles that mainly determines the exact timing of the epidemic within this window. The susceptible fraction of the population increases between consecutive epidemics as a result of a gradual reduction of antibody levels and antigenic shift of the pathogen (*38*). For an annually occurring epidemic, skipping of one season then leads to a larger fraction of susceptibles at the start of the next epidemic window. An analysis of the effective reproduction number within the context of an SIRS model (1) then predicts an earlier onset and peak of the epidemic (Fig. 4a), matching the epidemiological data (Fig. 1). This view on the temporal dynamics of the susceptible pool and the effective reproduction number is analogous to the notions of overcompensation and return time recently introduced to analyze the conditions for mutual invasibility and co-circulation of pathogens (*43*).

It is important to note that this shift in timing is not the result of an individual immunity debt or otherwise weaker immune system, since individual differences due to previous exposures to the pathogen is not part of the model. Instead any changes to the usual seasonal dynamics are the result of a loss of population-level immunity and a greater pool of susceptible hosts, not because of individually more severe infections.

The SIRS model also predicts that this increased pool of available hosts is depleted quickly during just one season, bringing the size of the susceptible fraction back to its usual pre-NPI level two seasons after the skipped season (Fig. 4a). The model thus predicts a relatively quick shift back to the normal timing and severity for seasonal respiratory epidemics, which indeed seems to be the case for influenza and RSV in Germany (Fig. 3).

Since a greater fraction of susceptibles leads to a higher effective reproduction number earlier in the season, the pathogen starts to spread earlier in its seasonal window and the predicted shift in epidemic timing is generally towards earlier in the season. But if the pathogen has been driven almost to extinction during the NPI season, even early-onset growth may not be enough to lead to a noticeable increase in disease incidence during the next season. In this case, repeated cryptic spread over one or more seasons is necessary before a noticable epidemic occurs again. This form of delayed resurgence has for example been described for bacterial respiratory infections with *Mycoplasma pneumoniae* (*44*).

A shift of 2-3 months in the seasonal timing of major respiratory diseases has profound practical implications. Doctors and hospitals may see a surge of cases when they do not usually expect them, leading to staffing and resource shortages. This is exacerbated by the shifted peaking of acute and severe respiratory cases during December in Germany, overlapping with the seasonal holidays. It also highlights the need to anticipate an earlier start to seasonal vaccination campaigns, such as against influenza, if the preceding season was unusually mild or entirely skipped. And epidemiologists and public health practitioners need to be aware that comparing epidemiological data from the current season to the same time point in previous seasons is not meaningful if a major disruption of the usual seasonal pattern has occurred.

One epidemic season has passed since the WHO declared the global COVID-19 pandemic over in May 2023. As we have seen from the data on the incidences of respiratory infections, the overall seasonal dynamics of respiratory infections is returning back to its pre-pandemic pattern. Seasonal forcing of transmission also plays a role for SARS-CoV-2 (*45*), but here the situation remains more complicated due to newly arising variants, rapidly waning immunity and very high effective reproduction number.

Despite almost the entire population of Germany having been in contact with either SARS-CoV-2 or a vaccine by mid-2022 (*46*), the inherently much larger transmissibility of SARS-CoV-2 makes it prone to spread earlier in the season than other respiratory diseases (cf. Fig S5e). This is especially true for the later variants of SARS-CoV-2 and for the time being we should thus expect a succession of early seasonal onset of COVID-19 in fall, followed by seasonal influenza and RSV closer to their usual time windows of late winter and early spring. This seasonal timing suggests that a combined vaccination campaign may not be ideal to reduce the number of cases.

While we have focused on the epidemiology in Germany, repeatable and consistent timing is a hallmark of seasonal respiratory disease epidemics in many temperate regions (*47*). At the same time there is evidence for shifts in epidemic timing worldwide, suggesting that our observations hold more generally.

The minimal SIRS model neglects evolutionary aspects of seasonal epidemics arising from the intertwined processes of waning immunity on the host side and antigenic variation on the pathogen side (*48, 49*). But the very repeatable timing of seasonal epidemics suggests that such processes do not generally lead to large-scale shifts in the timing and peak of epidemics. This either requires the emergence of a novel pathogen into largely immune-naive population or a major disruption of the usual seasonal dynamics, for example through widespread implementation of sufficiently strict NPIs.

Our analyis of epidemiological data and an SIRS model highlights that disruptions to the usual seasonal dynamics of respiratory infections not only affect the severity, but even more so the otherwise very consistent timing of seasonal epidemics. A simple SIRS model reproduces this shift across a wide range of scenarios, showing that such large scale patterns can be explained by the population level epidemiology of shifting susceptible and infective subpopulations. Beyond explaining the seasonal shift following the SARS-CoV-2 pandemic our analysis shows more generally that, within the exogenously determined window of epidemic potential, endogenous variables such as the availability of susceptibles are key for the exact timing of epidemics.

## Data Availability

All data is openly available online from the Robert-Koch-Institut (http://github.com/robert-koch-institut) and from the Federal Statistics Office of Germany (http://www-genesis.destatis.de).

## Data and materials availability

The epidemiological data is available from the Robert-Koch-Institut, http://github.com/robert-koch-institut, (*50,51*). Mortality data is available from the Federal Statistics Office of Germany, http://www-genesis.destatis. de, (*35*). A snapshot of the specific data used in our analysis together with the code used for the analysis of the data and the SIRS model is available at https://github.com/misieber.

## Supplementary materials

**Figure S1.**
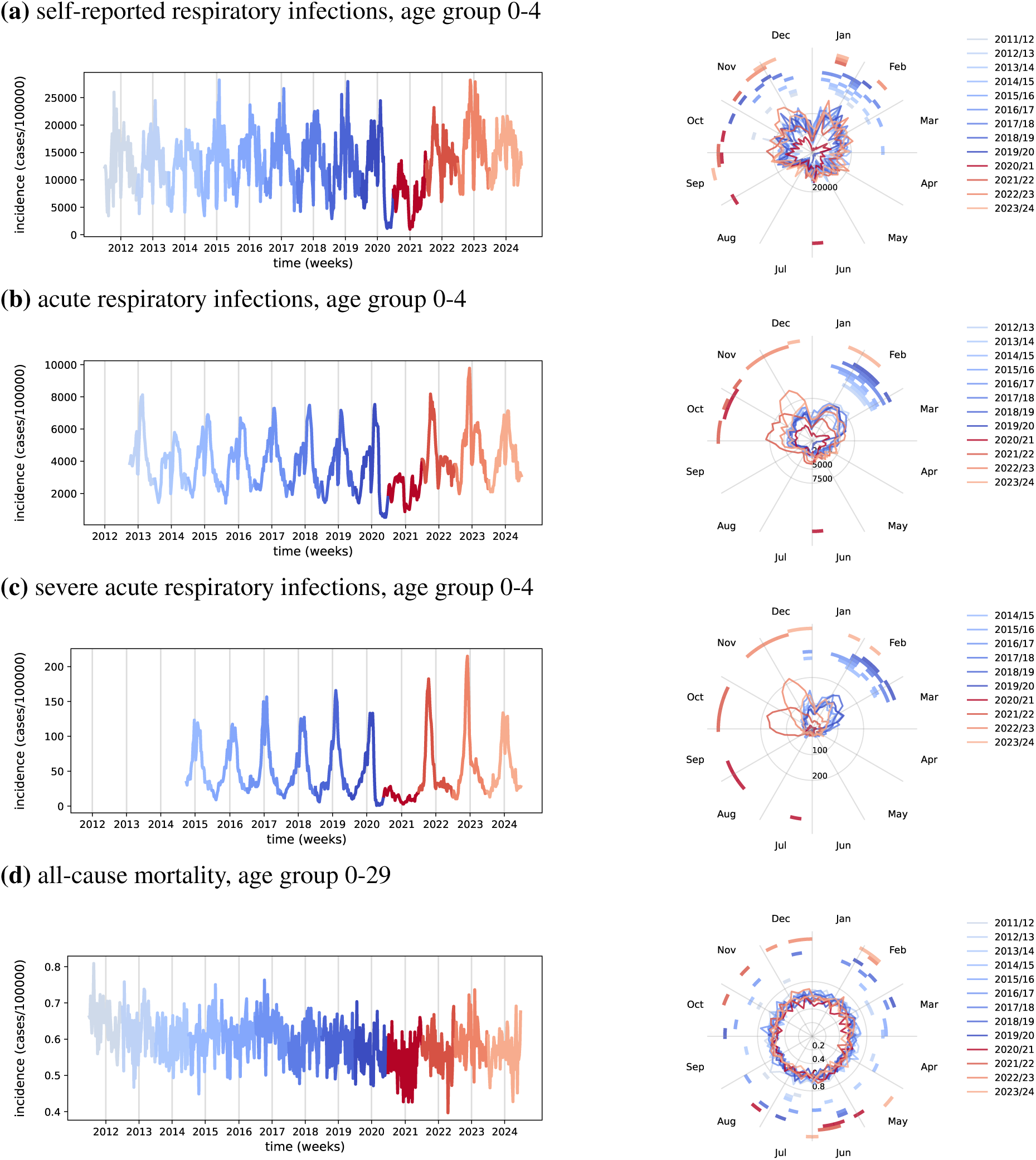
Weekly incidences of respiratory infections in Germany, young age group. Timeseries on the left and same data in a polar plot on the right to show the shift in peak respiratory infection season. (a) Self-reported, symptomatic respiratory infections (SRI). (b) Acute respiratory infections (ARI). (c) Hospitalized severe acute respiratory infections (SARI). (d) All-cause mortality as number of deaths per 100.000 per week.

**Figure S2.**
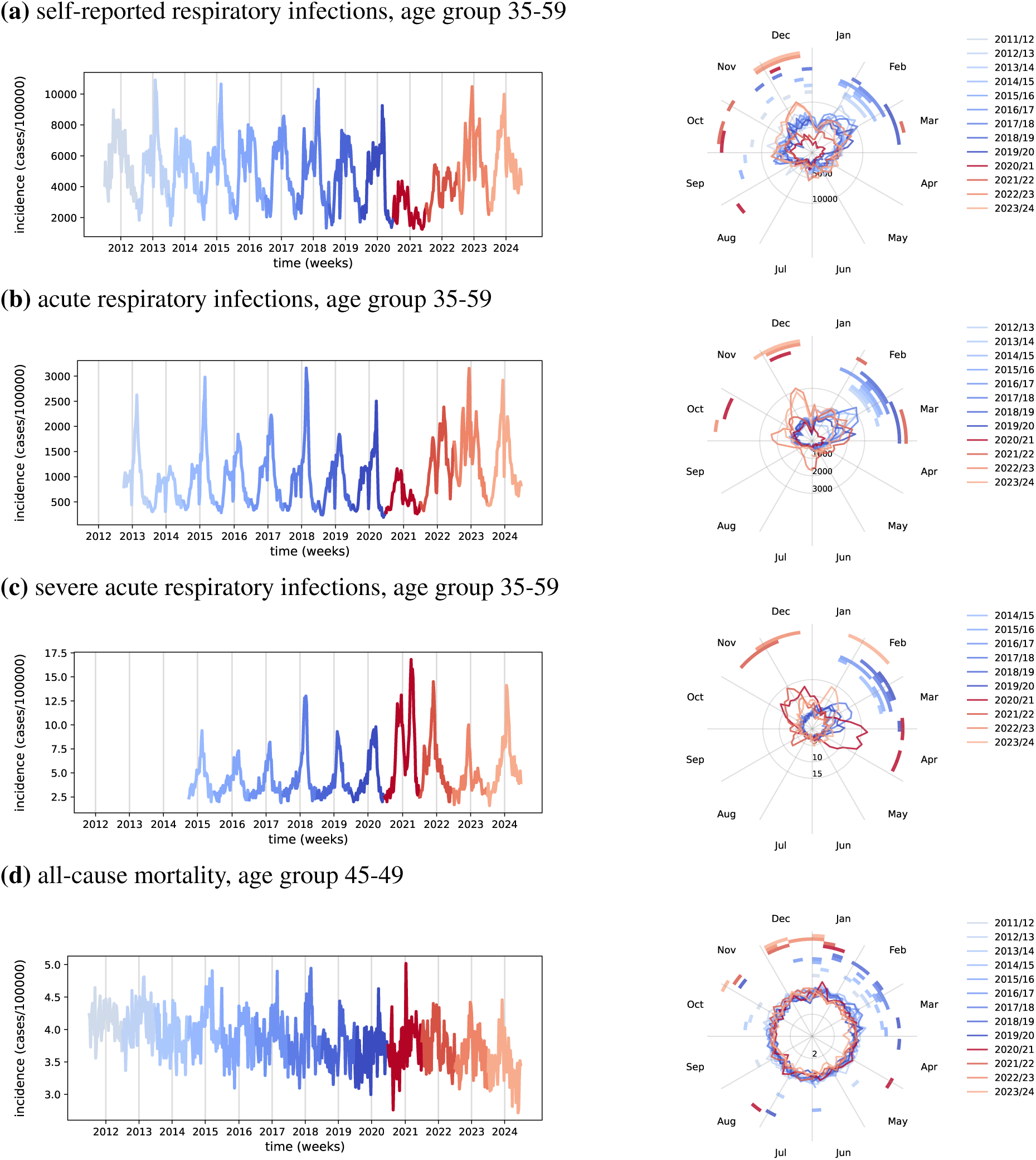
Weekly incidences of respiratory infections in Germany, intermediate age group. Timeseries on the left and same data in a polar plot on the right to show the shift in peak respiratory infection season. (a) Self-reported, symptomatic respiratory infections (SRI). (b) Acute respiratory infections (ARI). (c) Hospitalized severe acute respiratory infections (SARI). (d) All-cause mortality as number of deaths per 100.000 per week.

**Figure S3.**
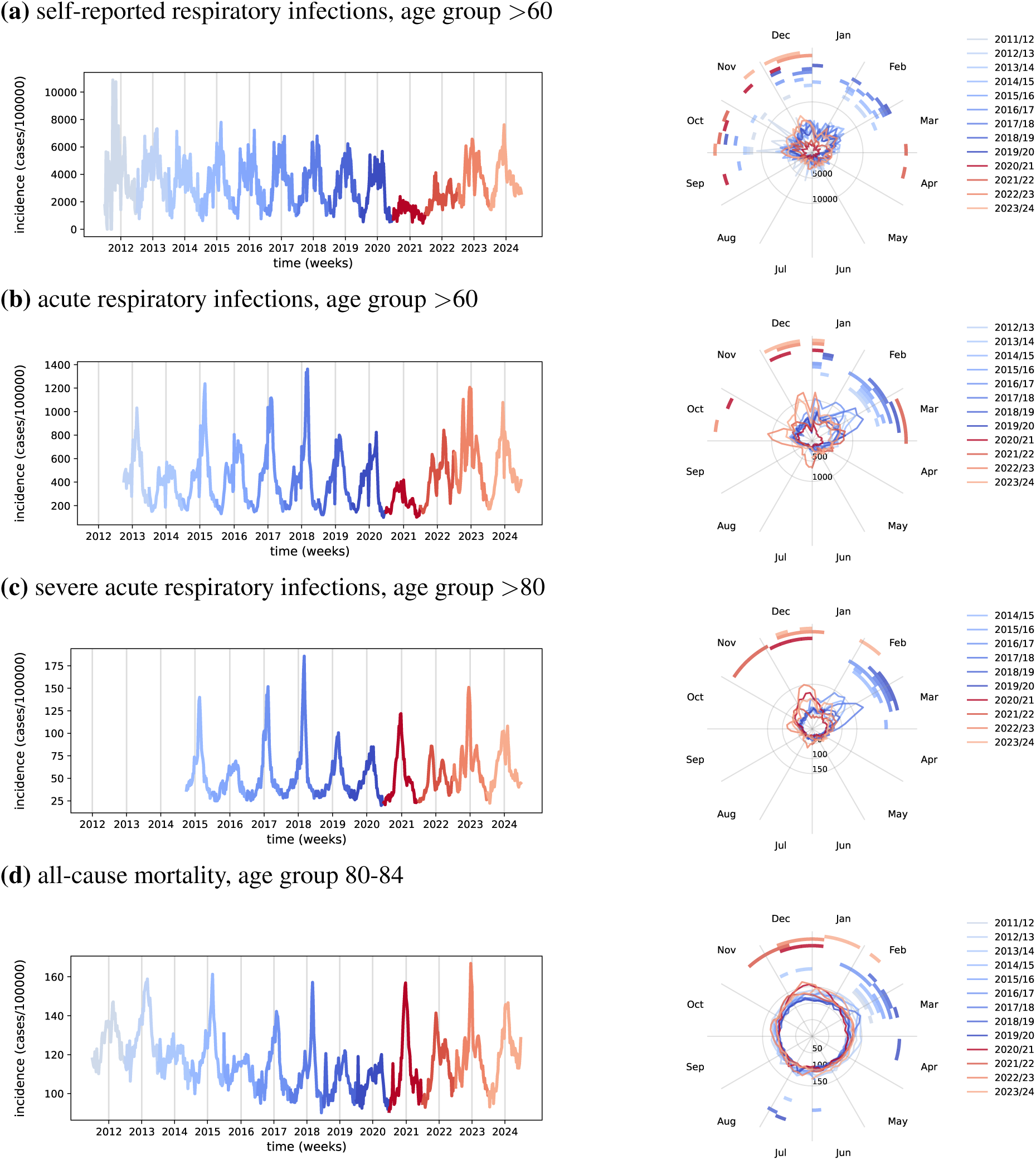
Weekly incidences of respiratory infections in Germany, old age group. Timeseries on the left and same data in a polar plot on the right to show the shift in peak respiratory infection season. (a) Self-reported, symptomatic respiratory infections (SRI). (b) Acute respiratory infections (ARI). (c) Hospitalized severe acute respiratory infections (SARI). (d) All-cause mortality as number of deaths per 100.000 per week.

**Figure S4.**
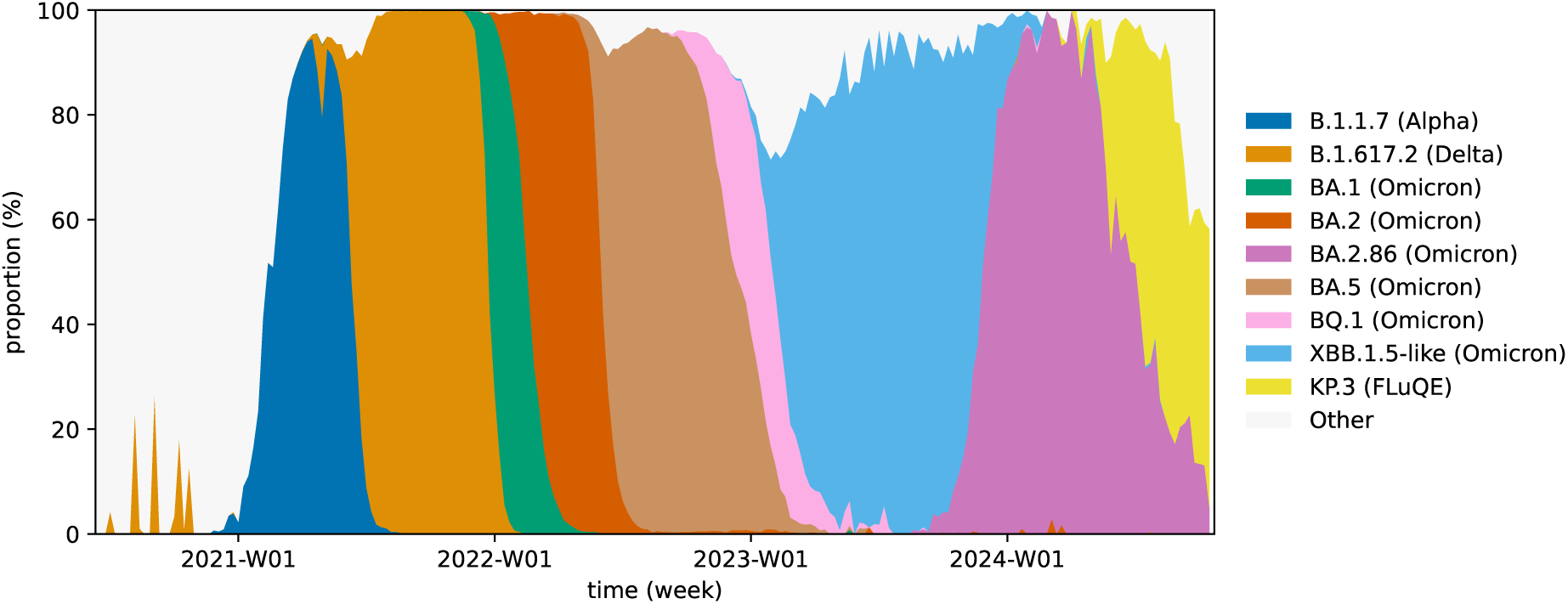
Timeline of major SARS-CoV-2 variants in Germany. Shown are only variants which reached a proportion of at least 25% at some point. Data source: European Centre for Disease Prevention and Control, http://github.com/EU-ECDC/Respiratory_viruses_weekly_data.

**Figure S5.**
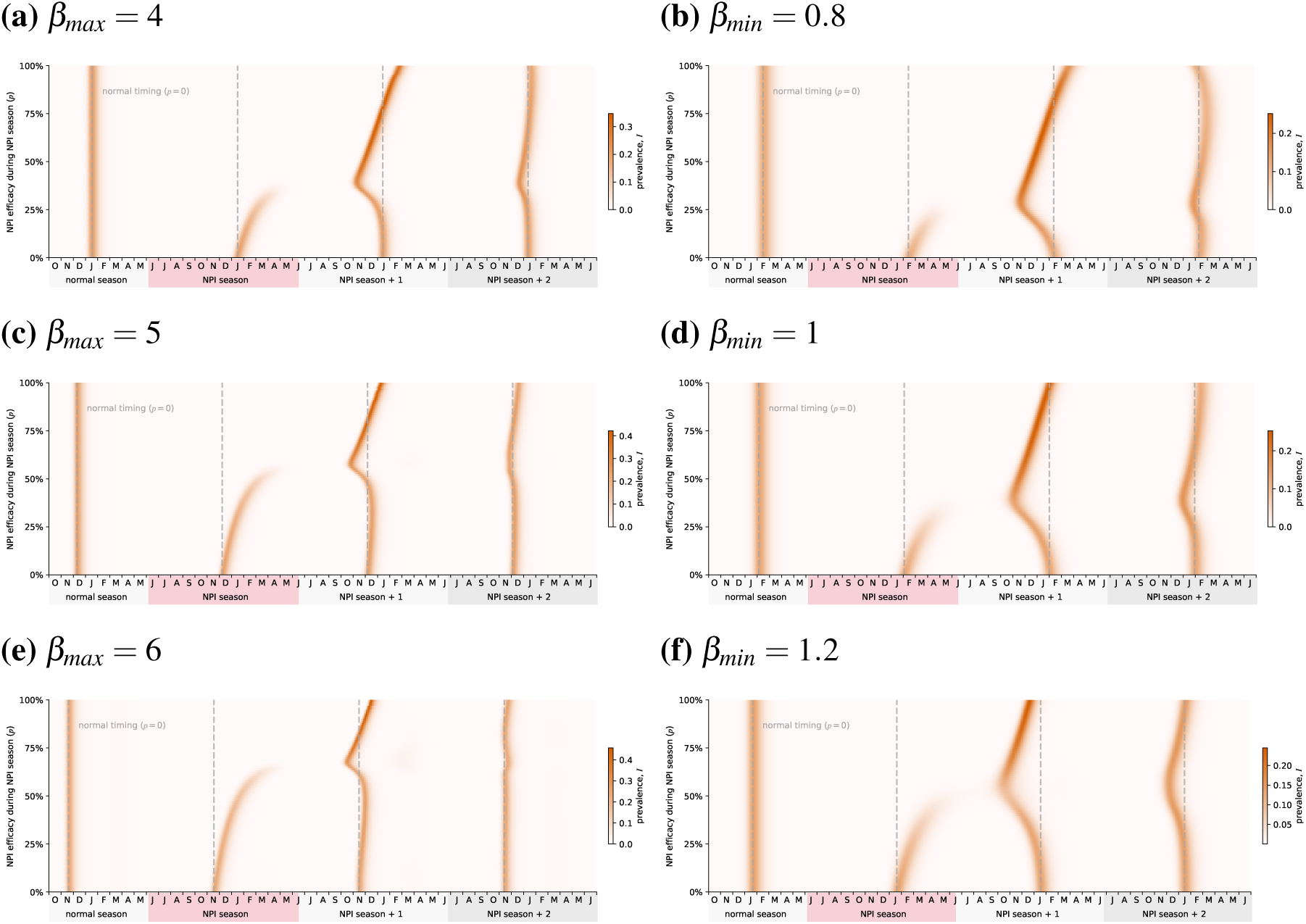
(a),(c),(e) In the absence of NPIs (*p* = 0), increasing the maximum transmission rate *β_max_*moves the seasonal peaks to an earlier timepoint, from late February for *β_max_* = 3 to November for *β_max_*= 6. An NPI season causes a similar disruption as described in the main text for *β_max_* = 3, but the effect is diminished at higher transmission rates as the usual seasonal peak gets closer to its earliest possible timing. (b),(d),(f) Increasing the minimum transmission rate *β_min_* also overall moves the seasonal peak forward, and exacerbates the effect of an NPI season.

**Figure S6.** Duration of immunity *ω*^−1^ = 10 weeks.

**Figure S7.**
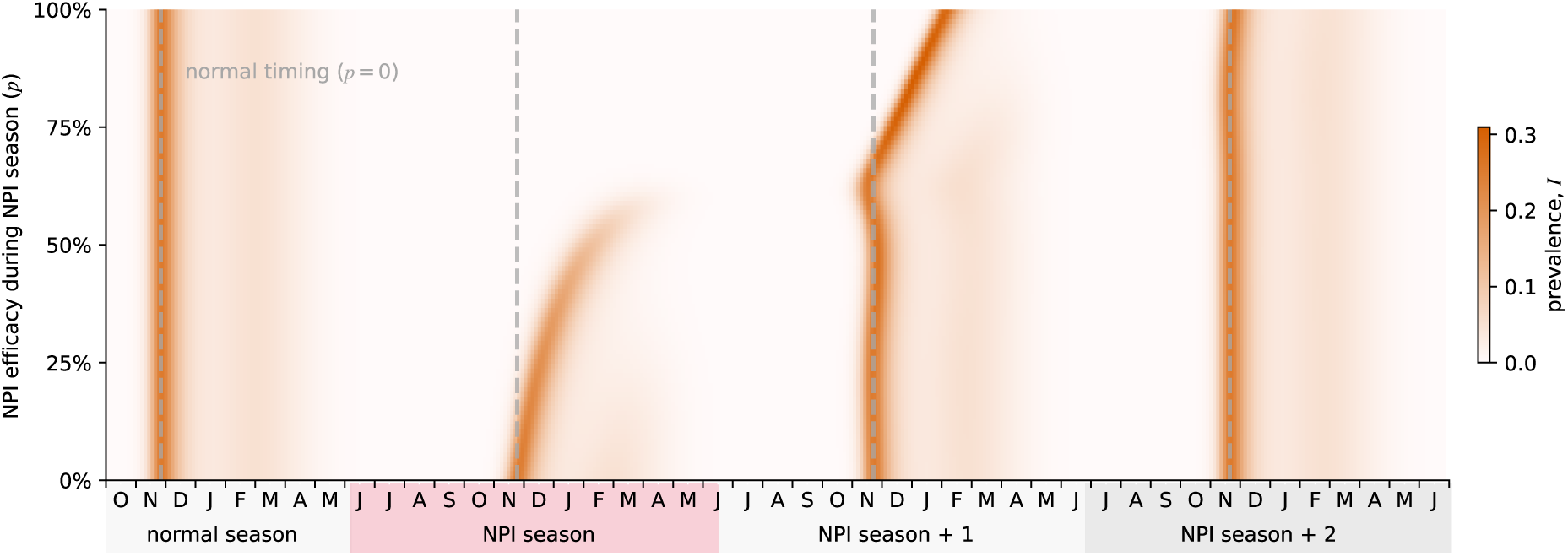
If immune waning happens much faster than the period of the underlying seasonal forcing (52 weeks), buildup of susceptibles is so fast that disruptions of the usual cycle have little effect, other than delaying the next season at very high NPI efficacies due to the disease being almost driven to extinction.

